# Changing patterns of SARS-CoV-2 infection through Delta and Omicron waves by vaccination status, previous infection and neighbourhood deprivation: A cohort analysis of 2.7M people

**DOI:** 10.1101/2022.04.05.22273169

**Authors:** Mark A. Green, Daniel J. Hungerford, David M. Hughes, Marta Garcia-Fiñana, Lance Turtle, Christopher Cheyne, Matthew Ashton, Gary Leeming, Malcolm G. Semple, Alex Singleton, Iain Buchan

## Abstract

**Objective:** To examine if SARS-CoV-2 infections vary by vaccination status, if an individual had previously tested positive and by neighbourhood socioeconomic deprivation across the Delta and Omicron epidemic waves of SARS-CoV-2.

**Design:** Cohort study using electronic health records

**Setting:** Cheshire and Merseyside, England (3^rd^ June 2021 to 1^st^ March 2022)

**Participants:** 2.7M residents

**Main Outcome measure:** Registered positive test for SARS-CoV-2

**Results:** Social inequalities in registered positive tests were dynamic during the study. Originally higher SARS-CoV-2 rates in the most socioeconomically deprived neighbourhoods changed to being higher in the least deprived neighbourhoods from the 1^st^ September 2021. While the introduction of Omicron initially reset inequalities, they continued to be dynamic and inconsistent. Individuals who were fully vaccinated (two doses) were associated with fewer registered positive tests (e.g., between 1^st^ September and 27^th^ November 2021: (i) individuals engaged in testing – Hazards Ratio (HR) = 0.48, 95% Confidence Intervals (CIs) = 0.47-0.50; (ii) individuals engaged with healthcare - HR = 0.34, 95% CIs = 0.33-0.34). Individuals with a previous registered positive test were also less likely to have a registered positive test (e.g., between 1^st^ September and 27^th^ November 2021: (i) individuals engaged in testing - HR = 0.16, 95% CIs = 0.15-0.18; (ii) individuals engaged with healthcare - HR = 0.14, 95% CIs = 0.13-0.16). However, Omicron is disrupting these associations due to immune escape resulting in smaller effect sizes for both measures.

**Conclusions:** Changing patterns of SARS-CoV-2 infections during the Delta and Omicron waves reveals a dynamic pandemic that continues to affect diverse communities in sometimes unexpected ways.

## Introduction

Vaccination is the cornerstone of preventing severe COVID-19 disease among individuals infected with the SARS-CoV-2 virus (1). Vaccines have also provided some protection from becoming infected with SARS-CoV-2, as has prior infection (2–4). Unvaccinated individuals are at higher risk of severe illness, hospitalisation or death from COVID-19 (4,5). There is a lack of evidence over how long either vaccine- or infection-acquired immunity to SARS-CoV-2 infection and COVID-19 disease may last for (6). Concerns over waning immunity (7,8), and immune escape with the Omicron variant (9), led to the introduction of ‘booster’ vaccination programmes in late 2021 (10). In addition to loss of biological protection, the risk behaviours of individuals change over time and may be influenced by feeling protected by vaccination or prior infection for longer than they actually are (11,12). Modelling of seasonal influenza vaccination programmes suggests that such behaviour changes can offset the effectiveness of vaccination programmes (13).

The COVID-19 pandemic has reinforced and amplified existing social inequalities in health. The number of infections, hospitalisations and deaths due to COVID-19 are disproportionally higher among residents of socioeconomically deprived neighbourhoods (14,15). Vaccination uptake is also lower among deprived populations (16). Assessing the importance of vaccine- and infection-acquired immunity are therefore social issues. However, current debates and evaluations of these issues largely ignore this social dimension. For example, estimates of vaccine effectiveness at reducing infections often present only unadjusted associations (17), which do not account for the differing levels of exposure to SARS-CoV-2 and vaccine uptake among different population and social groups.

The aim of this study is to examine if SARS-CoV-2 infections in England varied by vaccination status, if an individual had previously tested positive and by neighbourhood socioeconomic deprivation. We compare experiences during the epidemic curves of two SARS-CoV-2 variants: Delta and Omicron. The periods where these variants dominate infections represent an interesting case study due to high number of infections, high vaccine uptake, limited non-pharmaceutical interventions and changing public responses to national COVID-19 measures.

## Methodology

### Study design

Data were accessed from the Combined Intelligence for Population Health Action (CIPHA; www.cipha.nhs.uk) resource. CIPHA is a population health management data resource set up to support responses to COVID-19. CIPHA contains linked pseudonymised electronic health care records for 2,864,997 people. We restricted analyses to only people (n = 2,767,027) with a complete address who were resident in the integrated care region the CIPHA resource was set up to serve (Cheshire and Merseyside, England). We utilise data on our population (compiled from all people registered with a GP), linked to vaccination records and registered SARS-CoV-2 tests.

We select data to cover the following three time periods:

1. <u>Delta – 3rd June to 1st September 2021</u>: We define the start of the period as when Public Health England (now UK Health Security Agency) stated that the Delta variant was 99% of all infections (18).
2. <u>Delta – 1^st^ September to 27^th^ November 2021</u>: We select this period to cover the wave of infections associated with the new school year (starting 1^st^ September 2021) up to where the first case of Omicron was detected in England. The latter period was selected to focus our analyses on cases relating primarily to the Delta variant of SARS-CoV-2 to avoid any differences in risk of further infection or vaccine escape the Omicron variant may have.
3. <u>Omicron – 13^th^ December 2021 to 1^st^ March 2022</u>: We define the start of this period as when sequencing data suggested that most positive tests were for Omicron. The period is then up to the end of available data at the time of analysis.

For each period, we only include people who were alive up to the end of the period to minimise issues with immortal time bias.

### Outcome variable

The primary outcome variable was time to SARS-CoV-2 infection (registered positive test) during each period. Time was defined as when the test was taken rather than when it was processed. Positive cases are compiled from data feeds supplied by the UK Health Security Agency, who share all Pillar 1 (tests in care settings) and Pillar 2 (tests in the community) positive tests which are linked within CIPHA. Positive cases are identified using both lateral flow and polymerase chain reaction (PCR) tests.

### Explanatory variables

We focus on three key explanatory variables: COVID-19 vaccination status, previous SARS-CoV-2 infection and neighbourhood socioeconomic deprivation.

Vaccination status was defined as the number of doses (of any vaccine type combination e.g. BNT162b2 (Pfizer-BioNTech) and the ChAdOx1 nCoV-19 (Oxford-AstraZeneca)) an individual had received (0-3). We identified the number of first doses received two weeks before the start of each period, and one week prior for two or three doses, which we define as the time to receive immune protection (following other research (3,8)). The measure was then updated (i.e., time-varying) over time to account for people who received an additional vaccination during each study period.

Previous SARS-CoV-2 infection (binary) was defined as whether an individual had a registered positive test two weeks before the start of each period (19). The measure was held constant and not time varying. We defined this two-week period as the time to develop immune protection. Infections were selected based on the first positive test, and subsequent positive tests occurring more than 90 days apart (which we define as a further/subsequent infections). This definition follows established research elsewhere (8,19). We evaluated if this definition affected our results by introducing immortal time bias (i.e., some individuals could not test positive for parts of outcome periods if tested positive closer to the start period) and report these as a sensitivity analysis.

Neighbourhood socioeconomic deprivation was measured through matching individual’s residence to the 2019 Index of Multiple Deprivation (IMD) (20). The IMD is a multi-dimensional index of neighbourhood deprivation, based on seven weighted domains including income, employment, education and health. The IMD score is measured for Lower Super Output Areas (LSOAs) which are small zones representing neighbourhoods (∼1500 people). We also report analyses by IMD decile to aid interpretation.

### Control variables

We account for demographic factors sex (male or female) and age. Age was included as a categorical variable to account for non-linear dynamics and produced a better fitting model than a continuous measure. Age is an important factor for different risks in exposure to SARS-CoV-2, as well as to reflect that the vaccination programme was rolled out by age group. Ethnicity was included to account for inequalities in both exposure to SARS-CoV-2 and vaccination uptake. Broad ethnic groups were used: White, Asian, Black, Mixed and Other. We also include ‘prefer not to say / missing’ as a category in our models, since they account for a large proportion of records and this can account for any issues with this group being different in causal behaviours. Health status was included to account for differences in behaviours, where people with long-term health conditions may’shield’ or minimise social contacts. We define health status (comorbidity) as if individuals had a registered Expanded Diagnosis Clusters codes (yes or no). Codes represent diseases, symptoms or conditions that are treated in ambulatory and inpatient hospital settings. Finally, we also adjusted for differences in testing dynamics by accounting for whether an individual had registered a negative test in the previous month.

### Statistical analyses

We found evidence of inequalities in registered test behaviours (Appendix Table A). To minimise this potential bias in our regression analyses, we focus our analyses on two cohorts. First, we select only individuals who reported a negative test in the month prior to each time period as a proxy for being engaged in tested. This is similar to a ‘test-negative’ study design which have been used for studying vaccine effectiveness (21). Second, we analysed individuals who had received an influenza vaccine within a year of each time period as a proxy of being engaged in healthcare (i.e., likely to register a test even if unvaccinated and not disengaged with health care) (22). For the Omicron period, we extended this time frame to 1^st^ September 2020 to fully capture the previous year’s influenza vaccination campaign. While our main models use all individuals, in a sensitivity analysis we restrict this population to just people aged 65 years and over as they are the focus of the UK influenza vaccination programme. Matching methods were also investigated for balancing populations across our exposure variables, but did not significantly alter the models and are not discussed here. We also report analyses for all residents of Cheshire and Merseyside as a sensitivity analysis.

Descriptive statistics and visualisations were produced to summarise our data and identify key trends. Cox regression models were then used to predict the associations between our explanatory and control variables to our outcome variable (time to registered positive test). Hazard ratios and 95% confidence intervals were estimated from these models to summarise associations. Participants with missing data (n=101) were excluded from analyses (other than for ethnicity which we adjust for). Interaction effects for vaccine status and previous infection were tested, but not included in the results since they did not improve the model fit. We also stratified analyses by 10-year age group. This was to capture dynamics between children/adolescents and adults which will each have different modes of transmissions, risks and vaccination access (23).

#### Patient and Public Involvement

No patients and the public were involved in this piece of research.

## Results

Table 1 presents the descriptive characteristics of our cohort. Figure 1 presents trends in registered positive cases for all residents since the start of the pandemic to contextualise our three periods. The number of cases remains high during both Delta periods compared to previous waves. Omicron saw large growth in cases (10.5% of all residents registered a positive test; more than twice as high as both Delta periods) from new infections and the emergence of subsequent infections that almost reach the levels of new infections during the two Delta periods. We estimate that 11.4% of positive tests during the Omicron period were subsequent positive tests (in the other two periods, this figure was <1%). In particular, incidence of further infections were roughly twice as high in the most deprived compared to least deprived areas (Appendix Figure A). Percentage of people with registered positive tests across our exposure variables are described in Appendix Table B.

**Table 1:**
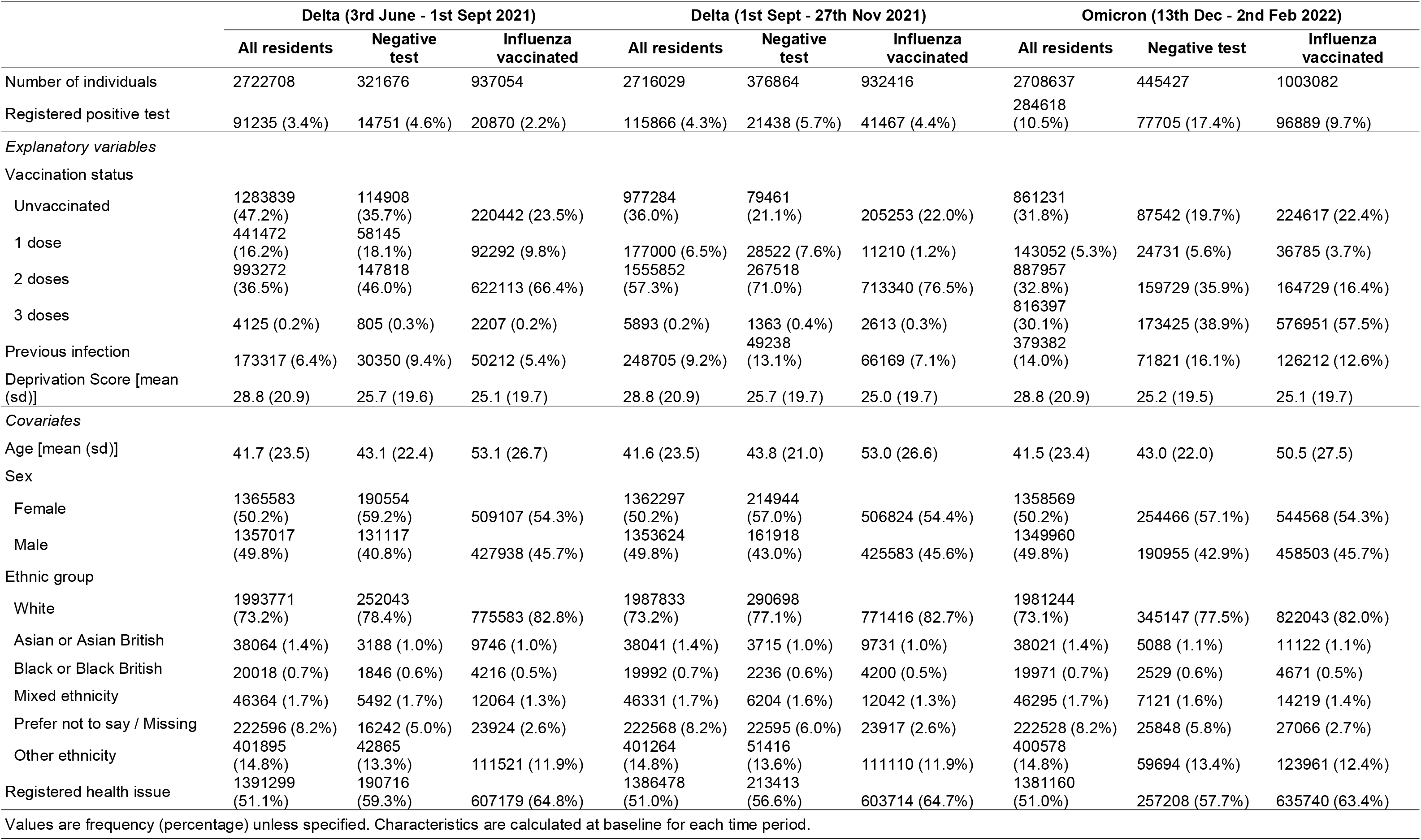
Sample characteristics for each period. Note: Values are frequency counts (percentage) unless specified.

**Figure 1:**
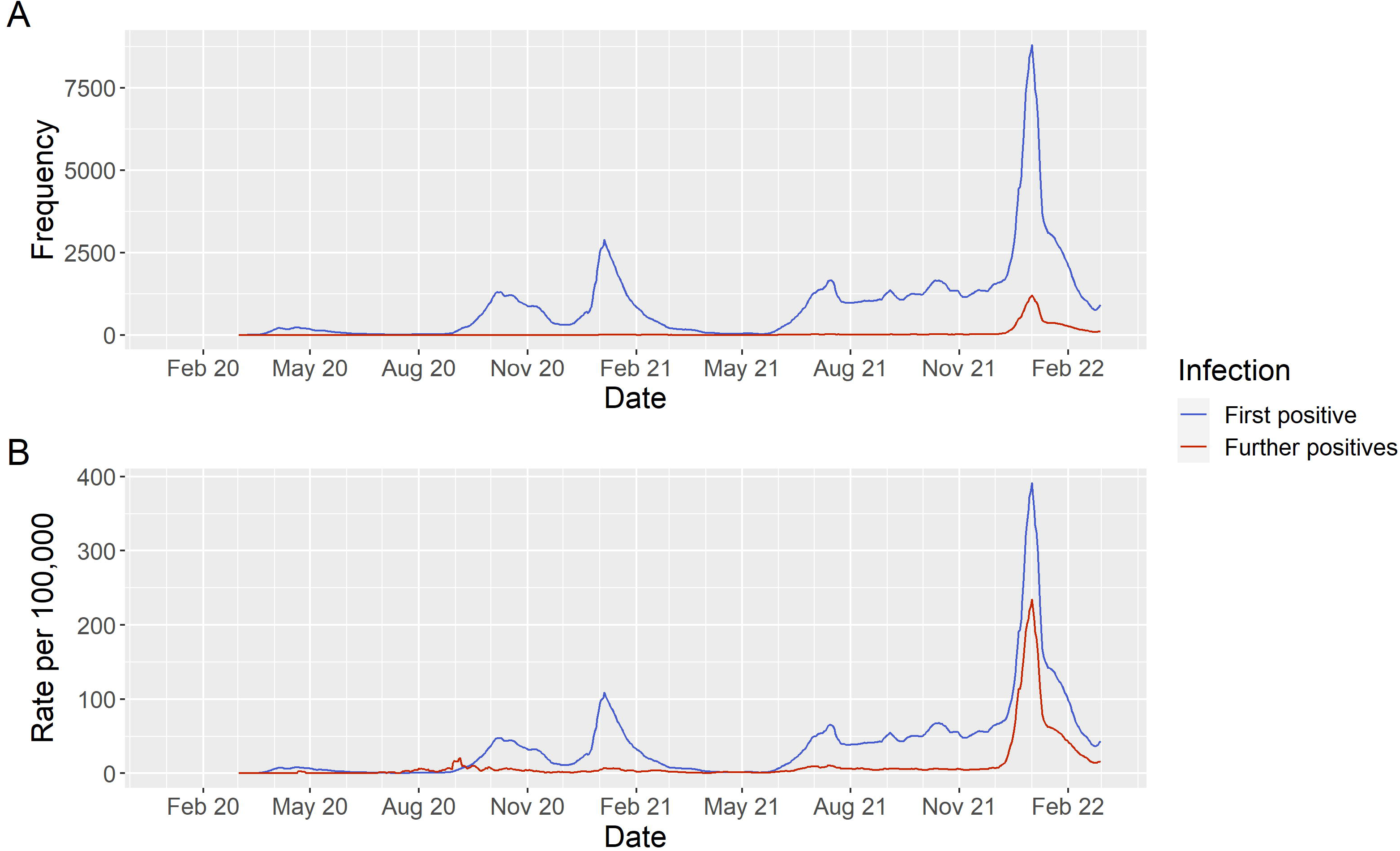
Seven day moving average for registered positive tests for all residents in Cheshire and Merseyside (England) by whether it was an individual’s first registered positive test (new infection) or further/subsequent positive test. A = Total number of cases, B = Total number of cases per 100,000 population.

Tables 2 (individuals engaged in testing) and 3 (individuals engaged in healthcare) presents findings from a series of Cox regression models predicting factors associated with time to registered positive test. There is agreement in associations across both model types for the two Delta periods, with less consistent findings for the Omicron period.

**Table 2:** Results for a Cox regression predicting time to a registered positive test for individuals who had registered a negative lateral flow test within a month of the time period start date (as a proxy for testing engaged).

|  | Delta (3rd June - 1st Sept 2021) |  |  | Delta (1st Sept - 27th Nov 2021) |  |  | Omicron (13th Dec - 2nd Feb 2022) |  |  |
| --- | --- | --- | --- | --- | --- | --- | --- | --- | --- |
|  | HR | LCI | UCI | HR | LCI | UCI | HR | LCI | UCI |
| *Unadjusted* |  |  |  |  |  |  |  |  |  |
| Unvaccinated | Reference |  |  | Reference |  |  | Reference |  |  |
| 1 dose | 0.7002 | 0.6620 | 0.7407 | 0.4728 | 0.4412 | 0.5067 | 1.1642 | 1.1119 | 1.2190 |
| 2 doses | 0.4049 | 0.3904 | 0.4198 | 0.4819 | 0.4678 | 0.4964 | 1.6575 | 1.6136 | 1.7026 |
| 3 doses | 0.4077 | 0.2791 | 0.5957 | 0.2370 | 0.1890 | 0.2971 | 0.8649 | 0.8420 | 0.8885 |
| Previous infection | 0.2455 | 0.2193 | 0.2748 | 0.1611 | 0.1462 | 0.1774 | 0.7342 | 0.7127 | 0.7563 |
| Deprivation Score | 1.0049 | 1.0041 | 1.0056 | 0.9951 | 0.9944 | 0.9959 | 1.0045 | 1.0041 | 1.0049 |
| *Adjusted\** |  |  |  |  |  |  |  |  |  |
| Unvaccinated | Reference |  |  | Reference |  |  | Reference |  |  |
| 1 dose | 0.6434 | 0.6030 | 0.6866 | 0.6808 | 0.6332 | 0.7320 | 1.0441 | 0.9935 | 1.0973 |
| 2 doses | 0.5336 | 0.5078 | 0.5606 | 0.6558 | 0.6284 | 0.6843 | 1.2998 | 1.2581 | 1.3429 |
| 3 doses | 0.5202 | 0.3550 | 0.7625 | 0.3296 | 0.2636 | 0.4121 | 0.9292 | 0.8963 | 0.9633 |
| Previous infection | 0.2310 | 0.2060 | 0.2591 | 0.1522 | 0.1382 | 0.1675 | 0.6092 | 0.5913 | 0.6276 |
| Deprivation Score | 1.0020 | 1.0012 | 1.0028 | 0.9961 | 0.9954 | 0.9968 | 1.0015 | 1.0011 | 1.0020 |
| Definitions: HR = Hazard Ratio, LCI = 95% Lower Confidence Interval, UCI = 95% Upper Confidence Interval |  |  |  |  |  |  |  |  |  |
| Note: Deprivation score is numerical, with increasing values representing higher levels of deprivation |  |  |  |  |  |  |  |  |  |
| * Adjusted for age (10-year age bands), sex, ethnicity, long-term illness, time varying vaccination status (with an interaction to time), previous infection status (and interaction to time), and 2019 Index of Multiple Deprivation score |  |  |  |  |  |  |  |  |  |

Unadjusted associations for both Delta waves showed that people who were vaccinated had lower likelihoods of registered positive test for SARS-CoV-2. For instance, in individuals engaged in testing we estimate that people who were fully vaccinated (2 doses) were 60% (Hazard Ratio (HR) = 0.40, 95% Confidence Intervals (CIs) = 0.39-0.42) and 52% (HR = 0.48, 95% CIs = 0.47-0.50) less likely to have a registered positive test in the first and second Delta waves respectively compared to unvaccinated people. In individuals engaged in healthcare, we estimate a larger effect size with individuals who were fully vaccinated being 63% (HR = 0.37, 95% CIs = 0.36-0.39) and 66% (HR = 0.34, 95% CIs = 0.33-0.34) less likely to have a registered positive test in the first and second Delta waves respectively compared to unvaccinated people. After adjusting for other demographic and social factors that may affect exposure to the virus, the strength of associations reduced but remained negatively associated (i.e., 95% confidence intervals did not cross 1). In the second Delta wave (1^st^ September to 27^th^ November 2021), we observed a stronger protective effect in people who had received 3 doses in both models (i.e., fully vaccinated and ‘boosted’).

Associations during the Omicron period were different to the previous Delta periods and varied between models. For both models, unadjusted associations suggested positive associations in 1 or 2 doses, and a negative association for 3 doses (both compared to unvaccinated populations). For example, individuals engaged in testing with three doses were 14% less likely (HR = 0.86, 95% CIs = 0.84-0.89) and individuals who were healthcare engaged were 23% less likely (HR = 0.77, 95% CIs = 0.75-0.78). In adjusted models, associations were largely attenuated in the testing engaged model. However, for people who were healthcare engaged, we find negative associations for all levels of vaccination status suggesting that following adjustment for known risk factors that may affect exposure to SARS-CoV-2, vaccinated healthcare engaged individuals were less likely to have a registered positive test.

People with a previous registered positive test had lower likelihood of having a registered positive test in each period across both models. Unadjusted effect sizes were large. For example, between 1^st^ September and 27^th^ November 2021 (Delta) we estimate that individuals with had a previous registered positive test were 84% (testing engaged model HR = 0.16, 95% CIs = 0.15-0.18; Table 2) and 86% (healthcare engaged model HR = 0.14, 95% CIs = 0.13-0.16; Table 3) less likely to have tested positive than compared to those who had not. Associations were consistent following adjusting for other covariates. The unadjusted effect size was smaller in the Omicron period (testing engaged model HR = 0.73, 95% CIs = 0.71-0.76; healthcare engaged model HR = 0.58, 95% CIs = 0.56-0.60), although effect sizes strengthened upon adjustment. Sensitivity analyses suggested that these associations remained consistent following assessing if our measure was affected by immortal time bias (Appendix Table C).

**Table 3:** Full model results for a Cox regression predicting time to a registered positive test for individuals who had received an influenza vaccination within a year of the time period start date (as a proxy for healthcare engaged).

|  | Delta (3rd June - 1st Sept 2021) |  |  | Delta (1st Sept - 27th Nov 2021) |  |  | Omicron (13th Dec - 2nd Feb 2022) |  |  |
| --- | --- | --- | --- | --- | --- | --- | --- | --- | --- |
|  | HR | LCI | UCI | HR | LCI | UCI | HR | LCI | UCI |
| *Unadjusted* |  |  |  |  |  |  |  |  |  |
| Unvaccinated | Reference |  |  | Reference |  |  | Reference |  |  |
| 1 dose | 0.6936 | 0.6527 | 0.7371 | 0.3627 | 0.3254 | 0.4043 | 1.2674 | 1.2142 | 1.3228 |
| 2 doses | 0.3747 | 0.3641 | 0.3855 | 0.3372 | 0.3304 | 0.3442 | 1.3909 | 1.3535 | 1.4293 |
| 3 doses | 0.4349 | 0.3212 | 0.5889 | 0.1277 | 0.1062 | 0.1535 | 0.7660 | 0.7500 | 0.7823 |
| Previous infection | 0.1935 | 0.1689 | 0.2217 | 0.1440 | 0.1302 | 0.1593 | 0.5833 | 0.5643 | 0.6030 |
| Deprivation Score | 1.0061 | 1.0054 | 1.0067 | 0.9951 | 0.9946 | 0.9956 | 1.0009 | 1.0005 | 1.0013 |
| *Adjusted\** |  |  |  |  |  |  |  |  |  |
| Unvaccinated | Reference |  |  | Reference |  |  | Reference |  |  |
| 1 dose | 0.6634 | 0.6085 | 0.7233 | 0.5291 | 0.4691 | 0.5966 | 0.9770 | 0.9298 | 1.0265 |
| 2 doses | 0.5504 | 0.5139 | 0.5894 | 0.6001 | 0.5659 | 0.6364 | 1.1499 | 1.0914 | 1.2116 |
| 3 doses | 0.6314 | 0.4622 | 0.8626 | 0.2824 | 0.2355 | 0.3386 | 0.9421 | 0.8948 | 0.9919 |
| Previous infection | 0.1670 | 0.1456 | 0.1915 | 0.1146 | 0.1036 | 0.1266 | 0.4508 | 0.4359 | 0.4661 |
| Deprivation Score | 1.0031 | 1.0024 | 1.0038 | 0.9925 | 0.9920 | 0.9930 | 0.9992 | 0.9987 | 0.9996 |

Definitions: HR = Hazard Ratio, LCI = 95% Lower Confidence Interval, UCI = 95% Upper Confidence Interval
Note: Deprivation score is numerical, with increasing values representing higher levels of deprivation
\* Adjusted for age (10-year age bands), sex, ethnicity, long-term illness, number of tests in previous month, time varying vaccination status (with an interaction to time), previous infection status (and interaction to time), and 2019 Index of Multiple Deprivation score

The associations for neighbourhood deprivation vary across each time period. In the first period (Delta – 3^rd^ June to 1^st^ September 2021), we estimate positive associations in both models indicating that individuals in more deprived areas were more likely to have a registered positive test. To aid interpretation of this effect, we also estimated a model using national decile of deprivation (Appendix Tables D and E). Individuals engaged in testing who resided in the least deprived decile were 24% less likely (HR = 0.76, 95% CIs = 0.72-0.81) and individuals who were healthcare engaged were 33% less likely (HR = 0.67, 95% CIs = 0.63-0.70), both compared to people in the most deprived decile.

In the second Delta period (1^st^ September to 27^th^ November 2021), the direction of the association was negative suggesting that as areas become more deprived, registered positive tests decreased. Individuals engaged in testing who resided in the least deprived decile were 37% more likely (HR = 1.37, 95% CIs = 1.30-1.44) and individuals who were healthcare engaged were 37% more likely (HR = 1.37, 95% CIs = 1.32-1.42), both compared to people in the most deprived decile. (Appendix Tables D and E). Age-stratified models suggest that the reversal of social inequalities appears to be driven by cases in children and older adults (Appendix Figure B).

In the Omicron period (13^th^ December 2021 to 28^th^ February 2022), associations for deprivation showed diverging patterns across our models. Associations were positive in the testing engaged model (Table 2) and negative following adjustment in the healthcare engaged model (Table 3). This reflects the complexity in identifying associations over this period, where deprived and less deprived communities had the highest rates of registered positive tests at different points (Figure 2). Initially incidence rates were higher in the least deprived decile, with trends reversing due to a larger peak of infections in the most deprived decile post-Christmas. By the end of the period, social inequalities had reversed again with more positive tests in the least deprived decile. For subsequent infections, the social gradient is more distinct with higher rates in the most deprived decile for most of the period before converging together.

**Figure 2:**
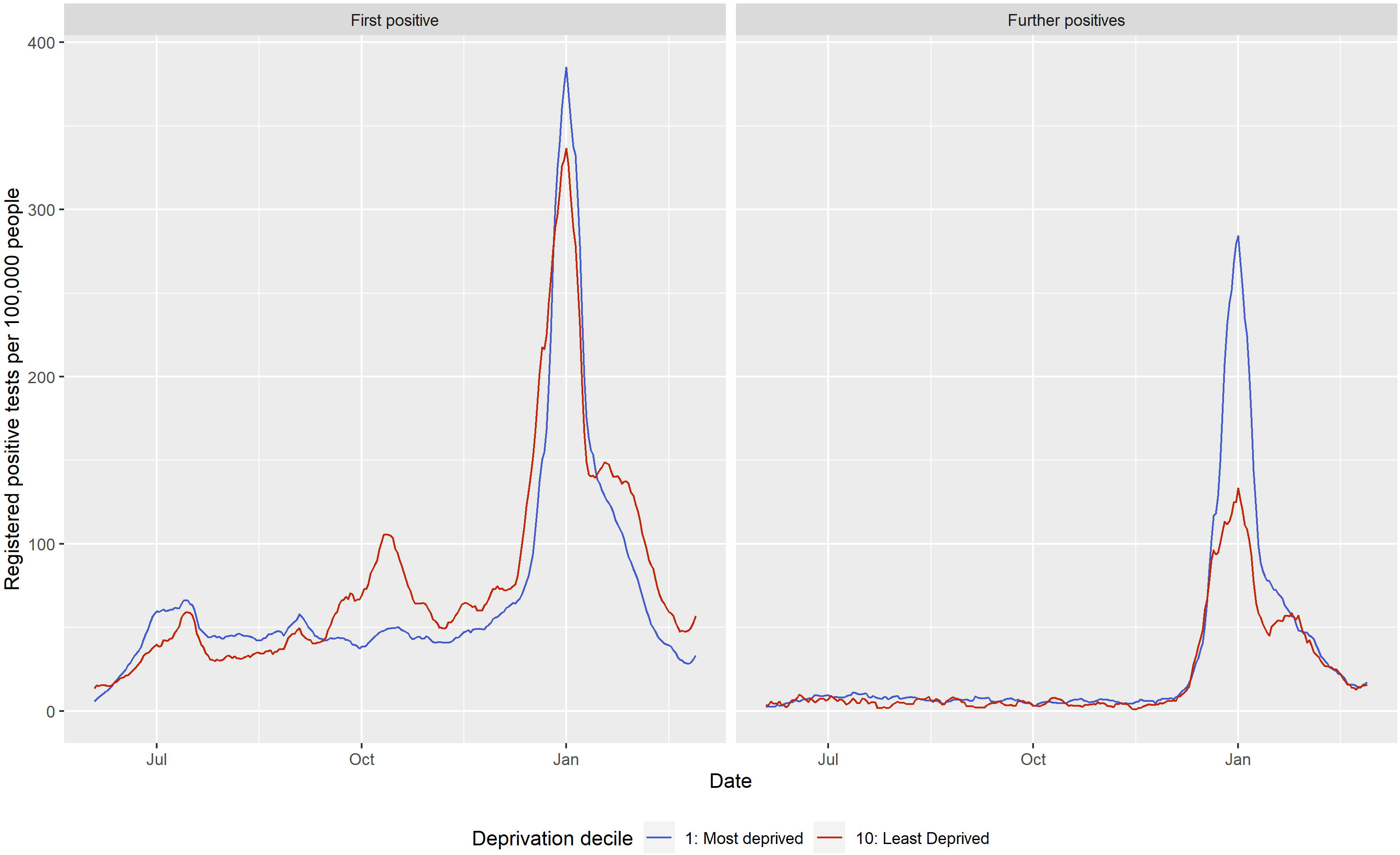
Comparison of seven day moving average for the number of residents in Cheshire and Merseyside per 100,000 people who registered a COVID-19 positive test for the most and least deprived deciles by whether it was an individual’s first registered positive test or a further/subsequent infection (3^rd^ June 2021 – 2^nd^ February 2022).

These results remain consistent when analysing all residents in Cheshire and Merseyside for all exposures other than for vaccination status (Appendix Table F), although any interpretation should be made cautiously due to the level of bias affecting associations. Similar associations were also reported when restricting the healthcare engaged individuals to only people aged 65 years and over, with some evidence of the changing vaccine associations earlier in the study period (Appendix Table G).

## Discussion

### Key results

Our study details the complex changes over time in who is being affected by the COVID-19 pandemic. While number of cases were high during the Delta waves, Omicron saw unprecedented numbers of cases with 10.5% of people in Cheshire and Merseyside having a registered positive test. Subsequent infections were identified in 11% of these tests, with rates higher in deprived areas. The types of people with registered positive tests continues to change. Initially, social inequalities were evident with registered positive tests were higher in the most deprived areas. Since 1^st^ September 2021, this has been less consistent with more registered positive tests in the least deprived areas (partly driven by patterns in children and adolescents). While there were fewer registered positive tests in vaccinated populations, this changed with Omicron. Finally, we find that people with a previous registered positive test are far less likely to have a subsequent registered positive test.

### Interpretation

Our study does not assess vaccine effectiveness or vaccine impact on SARS-CoV-2 infection or COVID-19 disease. Rather it describes the types of people with registered positive tests for SARS-CoV-2 during the Delta and Omicron waves, and the complexity in being able to tease out these associations. Our analyses demonstrated that most new infections in the Delta wave occurred among unvaccinated populations. This association, however, becomes less clear with the emergence of Omicron where in individuals engaged in testing we observe more registered positive tests in individuals who were fully vaccinated (but not for those who were boosted or in individuals engaged with healthcare). This is not to suggest that being vaccinated places people at greater risk of being infected. Causal explanations for this association may include behavioural changes, such as increased physical contacts and working outside the home following vaccination increasing exposure to the virus (11,12). Evidence in England suggests that while individuals did not change behaviours after being vaccinated, increasing population vaccination levels were associated to changes in risk-compensatory behaviours and social contacts (24). In addition, as most people get vaccinated or infected, the pool of unvaccinated people most susceptible to infection becomes smaller. It is plausible that this group is very different behaviourally and socially, and aversion to vaccination may translate to aversion to receiving or registering a test. For example, in the SIREN study where they test all individuals, they find fewer infections in vaccinated groups for each of our study periods (17), although the same study also has showed waning protection of vaccines in line with our findings (8). Additionally, their analyses do not adjust for other covariates that may explain exposure to the virus.

We find fewer registered positive tests in individuals with a previous positive test, with estimated effect sizes relatively larger than compared to vaccination status. This effect remains following adjusting demographic and social characteristics. The under-reporting of tests in individuals with a previous registered positive test may partly explain this difference. Immunity responses may also be different between vaccines and natural infections (6). A large protective effect in natural infection has been reported elsewhere (8,19). Our estimated effect size reduced during the Omicron period, suggesting that the variant may be more effective at immune escape when compared to Delta. This is further highlighted by the larger percentage of subsequent infections identified.

Our findings should not be interpreted as naturally acquired immunity being recommended over vaccination. It is difficult to fairly compare effects across different variable types to identify which is most important and our methods do not allow for this. The people in our study who were previously infected excludes those that died of COVID-19, and the benefits of safe and effective vaccines have been clearly demonstrated in reducing COVID-19 hospitalisations and deaths (1,4,5). However, our analyses might give some clues as to why England has not witnessed ‘herd immunity’ despite high levels of vaccination uptake.

From 1^st^ September 2021, we find evidence of SARS-CoV-2 infections being higher in the least socioeconomically deprived communities. This remains in contrast to trends in the earlier in the pandemic, which had seen consistently higher infections in the most deprived areas (15). The reversal of the social gradient in the Delta wave may be explained by several factors. One explanation may be the large protective effect of previous infections that we find. When combined with the concentration of infections in deprived areas in previous waves (15), this may have logically led to reduced population susceptibility to infections in more deprived communities during the Delta wave. When Omicron arrives it ‘resets’ these patterns since it is effectively a new serotype with immune escape (25), and the most deprived areas are affected more again. However, our analyses suggest that the reversal of the gradient was independent of previous infection and vaccination status of communities. A second explanation may regard the heterogeneity of social networks. The increasing socioeconomic segregation of who lives where (26) and school intakes (27), combined with low socioeconomic mixing and contact (28), may produce waves of infections that do not transfer between social groups and their closed networks. Our age stratified models suggest the reversal of social inequalities was strongest in children and adolescents, suggesting the importance of school dynamics in driving infections during Delta and Omicron (23). Finally, inequalities in testing dynamics may produce an artefactual effect. Lower propensity to get tested or to register a test in deprived areas may bias our observations (29).

### Limitations

SARS-CoV-2 infections were identified based on a registered positive test. There was limited community testing availability during the first wave of infections and access to lateral flow tests were not available until late 2020 (6^th^ November in Liverpool only, 3^rd^ December rest of region). These issues may lead to missed infections that would not be reported in our data resulting in under-counts for previous positive tests. Not all individuals may get tested, nor register their test, leading to undercounts of infections in our measures. We attempted to account for some of these issues by restricting analyses to individuals who had registered a negative test in the month before due to established inequalities in testing uptake (29). The impact of this can be seen by comparing the models to analyses for all residents (e.g., Table 2 and Appendix Table C). We also report significant inequalities in who reported negative tests across our exposure variables (Appendix Table A) which may bias underlying associations. The range of bias we are unable to observe shows how difficult it is to investigate these phenomena using routine data, so our results should not be over-interpreted.

Our analyses are descriptive and exploratory. We could not investigate the mechanisms that may underlie the associations we report (e.g., the processes that explain why social inequalities changed over time). We also are unable to account for all potential confounders or explanatory factors (e.g., number of social contacts). It is plausible that our model adjustment may not be able to disentangle the association between demographic/social factors and our exposures (including risk behaviours and testing frequency). Future research should test potential reasons behind relationships, as this will be key for designing effective interventions.

### Conclusion

Using linked NHS and public health testing records for 2.7M people in Cheshire and Merseyside, our study reveals the dynamic nature of SARS-CoV-2 infections through the Delta and Omicron waves. Socially patterned immunity by vaccination and prior infection is resulting in social flips in who is infected, producing complex socioeconomic inequalities. Finding ways to effectively communicate the risks in exposure and infections among populations based on the changing dynamics we uncover remains important. In the context of ‘living with COVID-19’ and the removal of most non-pharmaceutical interventions, our findings suggest that highly infectious SARS-CoV-2 variants will continue to spread unequally through society but not always in expected ways.

## Data Availability

Data are accessible via CIPHA (https://www.cipha.nhs.uk/). Requests can be made to the Data Access Committee for extracts of the larger-scale data which cannot be released openly due to information governance requirements. All analyses were undertaking using open source R statistical software and code is made openly available here https://github.com/markagreen/social_flip_COVID-19.

## Ethics approval

Ethical approval was granted by the University of Liverpool’s Research Ethics committee (REF: 10634).

## Transparency statement

The lead author affirms that the manuscript is an honest, accurate, and transparent account of the study being report; that no important aspects of the study have been omitted; and that any discrepancies from the study as originally planned (and, if relevant, registered) have been explained.

## Contributions

MAG, MGF and IB came up with the idea for the study. Study design was then refined by all authors. MAG, DMH and CC accessed the data, cleaned and prepared it for analysis. Analyses were led by MAG, with input from DJH, DMH, and MGF. MAG, DJH, DMH, MGF, LT and IB led on initially drafting the paper, with CC, GL, MA, AS and CS providing input on revising the paper. The corresponding author attests that all listed authors meet authorship criteria and that no others meeting the criteria have been omitted.

