## Appendix for "Changing patterns of SARS-CoV-2 infection through Delta and Omicron waves by vaccination status, previous infection and neighbourhood deprivation: A cohort analysis of 2.7M people"

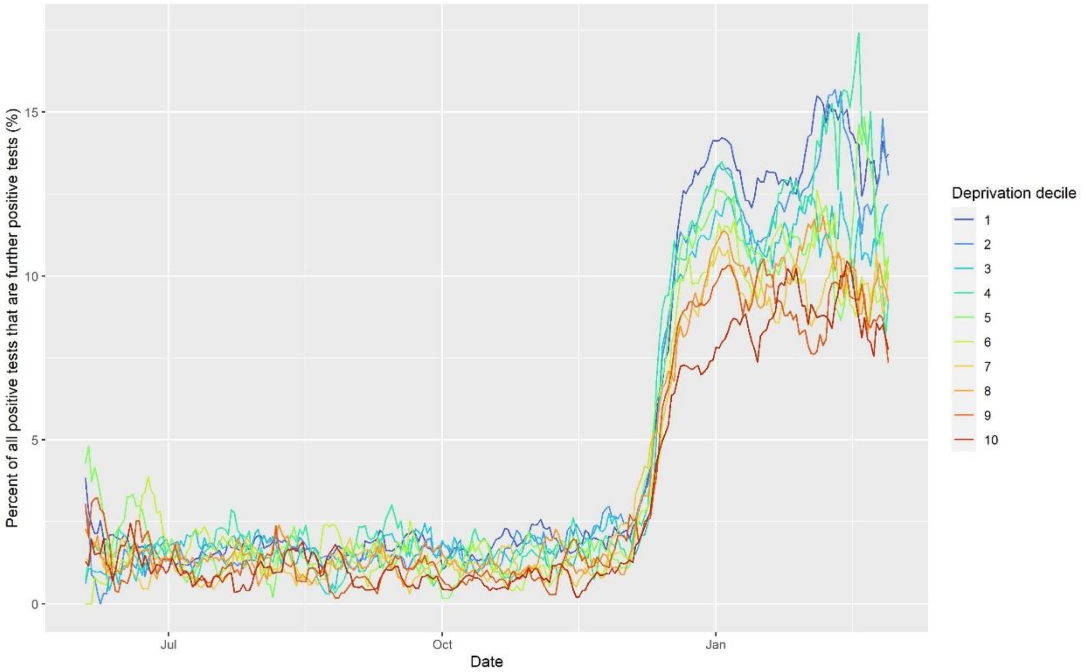


**Figure A: Seven day moving average for the percentage of all registered positive tests that were identified as a subsequent infection (>=2nd positive registered test more than 90 days apart) by decile of deprivation (3rd June 2021 – 2nd February 2022).**

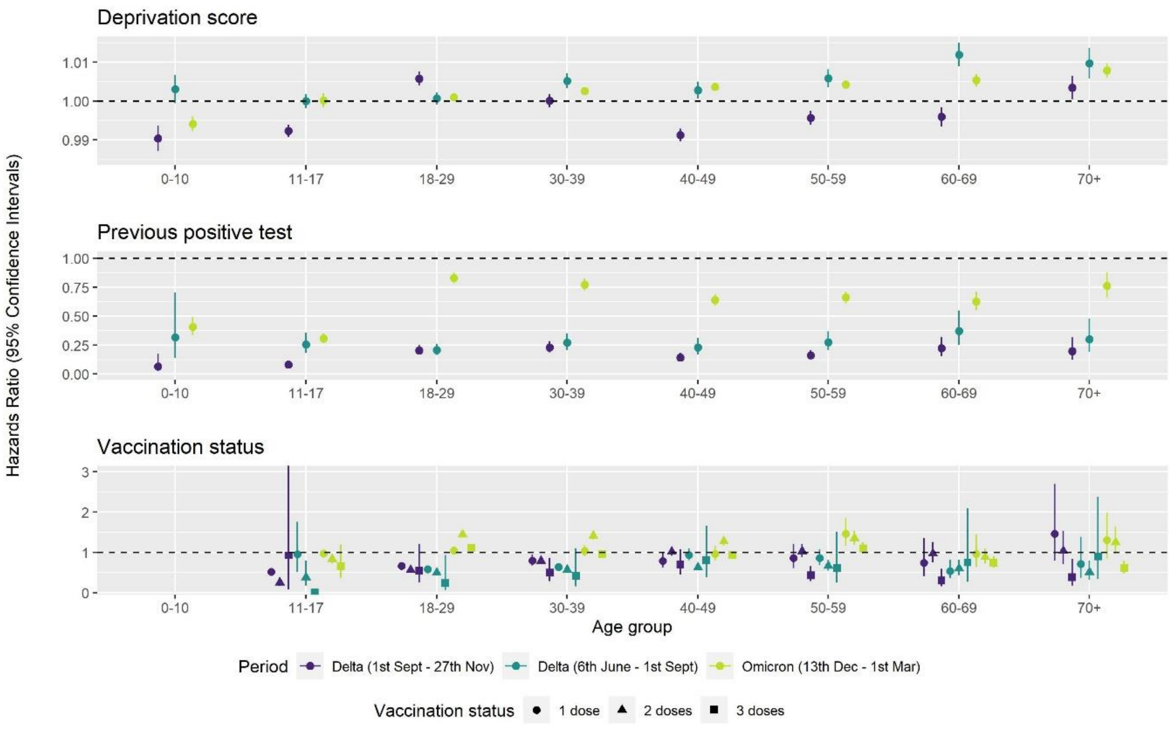


**Figure B: Age-stratified Cox regression models predicting time to registered positive test. Note: Coefficients for vaccination status for children aged 0-10 were not included, since due to low numbers model fit was not possible. Models are unadjusted for other covariates (model adjustment does not significantly change patterns).**

**Table A: Percentage of all participants in Cheshire and Merseyside who registered a negative COVID-19 test during each study period (i.e., within each outcome period).**

| | Delta (3rd June - 1st Sept 2021) | | | Delta (1st Sept - 27th Nov 2021) | | | Omicron (13th Dec - 2nd Feb 2022) | | |
|---|---|---|---|---|---|---|---|---|---|
| | All residents | Negative test | Influenza vaccinated | All residents | Negative test | Influenza vaccinated | All residents | Negative test | Influenza vaccinated |
| Unvaccinated | 24.4 | 66.4 | 28.5 | 27.9 | 60.3 | 38.2 | 19.9 | 47.8 | 27.0 |
| 1 dose | 31.1 | 73.8 | 33.3 | 33.1 | 60.9 | 34.4 | 41.3 | 69.5 | 57.2 |
| 2 doses | 32.2 | 76.0 | 30.6 | 36.0 | 70.4 | 34.8 | 30.4 | 60.3 | 31.8 |
| 3 doses | 40.9 | 84.0 | 38.3 | 44.3 | 77.8 | 42 | 34.3 | 72.2 | 33.1 |
| No previous infection | 27.7 | 71.7 | 29.8 | 32.1 | 67.2 | 34.9 | 27.4 | 62.2 | 31.3 |
| Previous infection | 38.1 | 77.6 | 40.2 | 41.1 | 69.9 | 43.9 | 37.4 | 67.1 | 40.3 |
| Most deprived decile (1) | 22.8 | 66.9 | 26.1 | 26.1 | 62.7 | 29.5 | 23.6 | 59.3 | 27.2 |
| Decile 2 | 26.0 | 70.6 | 28.7 | 30.4 | 66.3 | 33.3 | 26.7 | 61.5 | 30.1 |
| Decile 3 | 27.7 | 72.3 | 30.2 | 31.8 | 67.7 | 34.2 | 28.0 | 62.5 | 31.5 |
| Decile 4 | 28.3 | 72.7 | 31.0 | 32.8 | 67.5 | 35.9 | 28.4 | 63.0 | 31.9 |
| Decile 5 | 29.2 | 73.7 | 30.6 | 33.7 | 67.8 | 35.4 | 29.6 | 63.3 | 32.4 |
| Decile 6 | 30.9 | 73.5 | 31.7 | 35.8 | 69.3 | 37.1 | 31.3 | 64.3 | 33.8 |
| Decile 7 | 31.6 | 74.0 | 31.6 | 36.5 | 69.3 | 37.6 | 31.8 | 64.1 | 34.1 |
| Decile 8 | 32.1 | 74.3 | 32.1 | 37.2 | 69.2 | 38.0 | 32.3 | 64.6 | 34.4 |
| Decile 9 | 32.2 | 73.9 | 32.0 | 38.4 | 70.3 | 38.6 | 33.1 | 64.8 | 35.4 |
| Least deprived decile (10) | 34.1 | 76.2 | 33.6 | 40.1 | 70.9 | 40.5 | 34.5 | 65.8 | 37.1 |

Characteristics are calculated at baseline for each time period (and not time varying here).

**Table B: Percentage of all participants with a registered positive test by exposure variables.**

| | Delta (3rd June - 1st Sept 2021) | | | Delta (1st Sept - 27th Nov 2021) | | | Omicron (13th Dec - 28th Feb 2022) | | |
|---|---|---|---|---|---|---|---|---|---|
| | All residents | Negative test | Flu vaccinated | All residents | Negative test | Flu vaccinated | All residents | Negative test | Flu vaccinated |
| *Vaccination status* | | | | | | | | | |
| Unvaccinated | 4.5 | 7.0 | 4.1 | 5.9 | 10.1 | 10.0 | 8.6 | 18.4 | 13.6 |
| 1 dose | 3.1 | 4.0 | 2.4 | 2.7 | 3.6 | 2.9 | 12.6 | 18.8 | 14.6 |
| 2 doses | 1.9 | 2.9 | 1.5 | 3.5 | 4.7 | 2.9 | 13.8 | 19.7 | 10.6 |
| 3 doses | 2.5 | 3.0 | 1.8 | 3.9 | 4.3 | 3.4 | 8.6 | 14.7 | 7.5 |
| *Previous infection* | | | | | | | | | |
| No | 3.5 | 4.9 | 2.3 | 4.7 | 6.5 | 4.7 | 10.9 | 18.4 | 10.1 |
| Yes | 0.7 | 1.2 | 0.4 | 0.6 | 1.0 | 0.6 | 8.4 | 12.5 | 6.6 |
| *Deprivation level* | | | | | | | | | |
| Most deprived decile (1) | 3.6 | 5.3 | 2.7 | 3.6 | 4.9 | 3.8 | 9.9 | 18.8 | 9.4 |
| Decile 2 | 3.5 | 4.9 | 2.4 | 4.2 | 5.4 | 4.4 | 10.7 | 18.5 | 9.9 |
| Decile 3 | 3.4 | 4.6 | 2.3 | 4.3 | 5.6 | 4.4 | 10.9 | 17.9 | 10.1 |
| Decile 4 | 3.5 | 4.9 | 2.5 | 4.2 | 5.7 | 4.5 | 10.7 | 18.5 | 10.0 |
| Decile 5 | 3.4 | 4.4 | 2.2 | 4.3 | 5.5 | 4.3 | 10.7 | 17.8 | 9.6 |
| Decile 6 | 3.1 | 4.1 | 2.0 | 4.6 | 6.0 | 4.6 | 10.7 | 16.7 | 9.6 |
| Decile 7 | 3.1 | 4.3 | 2.0 | 4.6 | 5.9 | 4.3 | 10.6 | 16.6 | 9.6 |
| Decile 8 | 3.2 | 4.3 | 2.1 | 4.7 | 5.9 | 4.6 | 10.7 | 16.4 | 9.5 |
| Decile 9 | 3.0 | 3.9 | 1.9 | 5.0 | 6.5 | 5.0 | 10.8 | 16.3 | 9.5 |
| Least deprived decile (10) | 2.9 | 4.1 | 1.8 | 5.1 | 6.7 | 5.1 | 10.6 | 16.2 | 9.8 |

Note: Values are percentages. Characteristics are calculated at baseline for each time period.

**Table C: Model results when redefining the measure for previous positive test (i.e., excluding all individuals with a registered positive test within 90 days of the start of the period) to assess the extent of immortal time bias.**

| | **Delta (3rd June - 1st Sept 2021)** | | | **Delta (1st Sept - 27th Nov 2021)** | | | **Omicron (13th Dec - 2nd Feb 2022)** | | |
|---|---|---|---|---|---|---|---|---|---|
| | **HR** | **LCI** | **UCI** | **HR** | **LCI** | **UCI** | **HR** | **LCI** | **UCI** |
| Individuals who were registered a negative test | | | | | | | | | |
| *Unadjusted* | | | | | | | | | |
| Previous infection | 0.2323 | 0.2065 | 0.2612 | 0.1900 | 0.1718 | 0.2101 | 0.8719 | 0.8462 | 0.8984 |
| *Adjusted** | | | | | | | | | |
| Unvaccinated | Reference | | | Reference | | | Reference | | |
| 1 dose | 0.6428 | 0.6024 | 0.6860 | 0.6798 | 0.6322 | 0.7309 | 1.0244 | 0.9745 | 1.0768 |
| 2 doses | 0.5324 | 0.5067 | 0.5595 | 0.6470 | 0.6201 | 0.6752 | 1.2945 | 1.2532 | 1.3371 |
| 3 doses | 0.5201 | 0.3549 | 0.7622 | 0.3188 | 0.2546 | 0.3992 | 0.8888 | 0.8574 | 0.9212 |
| Previous infection | 0.2195 | 0.1948 | 0.2474 | 0.1884 | 0.1705 | 0.2083 | 0.7069 | 0.6860 | 0.7284 |
| Deprivation Score | 1.0020 | 1.0012 | 1.0028 | 0.9961 | 0.9954 | 0.9968 | 1.0011 | 1.0007 | 1.0016 |
| Individuals who were flu vaccinated | | | | | | | | | |
| *Unadjusted* | | | | | | | | | |
| Previous infection | 0.1835 | 0.1592 | 0.2115 | 0.1680 | 0.1508 | 0.1871 | 0.8229 | 0.7954 | 0.8513 |
| *Adjusted** | | | | | | | | | |
| Unvaccinated | Reference | | | Reference | | | Reference | | |
| 1 dose | 0.6621 | 0.6073 | 0.7219 | 0.5239 | 0.4645 | 0.5910 | 0.9284 | 0.8832 | 0.9758 |
| 2 doses | 0.5494 | 0.5130 | 0.5884 | 0.5959 | 0.5620 | 0.6319 | 1.1204 | 1.0643 | 1.1794 |
| 3 doses | 0.6313 | 0.4621 | 0.8624 | 0.2773 | 0.2312 | 0.3326 | 0.8858 | 0.8421 | 0.9318 |
| Previous infection | 0.1597 | 0.1384 | 0.1842 | 0.1531 | 0.1375 | 0.1704 | 0.6276 | 0.6066 | 0.6493 |
| Deprivation Score | 1.0031 | 1.0024 | 1.0038 | 0.9925 | 0.9920 | 0.9930 | 0.9987 | 0.9983 | 0.9991 |

Definitions: HR = Hazard Ratio, LCI = 95% Lower Confidence Interval, UCI = 95% Upper Confidence Interval

Note: Deprivation score is numerical, with increasing values representing higher levels of deprivation

* Adjusted for age (10-year age bands), sex, ethnicity, long-term illness, number of tests in previous month, time varying vaccination status (with an interaction to time), previous infection status (and interaction to time), and 2019 Index of Multiple Deprivation score

**Table D: Results for a Cox regression predicting time to a registered positive test by deprivation decile for individuals who had registered a negative lateral flow test within a month of the time period start date (as a proxy for testing engaged).**

| | Delta (3rd June - 1st Sept 2021) | | | Delta (1st Sept - 27th Nov 2021) | | | Omicron (13th Dec - 2nd Feb 2022) | | |
|---|---|---|---|---|---|---|---|---|---|
| | **HR** | **LCI** | **UCI** | **HR** | **LCI** | **UCI** | **HR** | **LCI** | **UCI** |
| *Unadjusted* | | | | | | | | | |
| Most deprived (1) | Reference | | | Reference | | | Reference | | |
| Decile 2 | 0.9217 | 0.8708 | 0.9757 | 1.0978 | 1.0414 | 1.1573 | 0.9487 | 0.9186 | 0.9797 |
| Decile 3 | 0.8636 | 0.8096 | 0.9212 | 1.1330 | 1.0702 | 1.1995 | 0.9224 | 0.8902 | 0.9556 |
| Decile 4 | 0.9274 | 0.8693 | 0.9894 | 1.1468 | 1.0819 | 1.2157 | 0.9532 | 0.9192 | 0.9885 |
| Decile 5 | 0.8538 | 0.7989 | 0.9125 | 1.1086 | 1.0457 | 1.1752 | 0.9206 | 0.8878 | 0.9545 |
| Decile 6 | 0.7785 | 0.7266 | 0.8341 | 1.1997 | 1.1331 | 1.2702 | 0.8262 | 0.7952 | 0.8583 |
| Decile 7 | 0.8030 | 0.7529 | 0.8564 | 1.2087 | 1.1445 | 1.2765 | 0.7959 | 0.7673 | 0.8255 |
| Decile 8 | 0.8085 | 0.7605 | 0.8594 | 1.1922 | 1.1317 | 1.2558 | 0.8191 | 0.7917 | 0.8476 |
| Decile 9 | 0.7307 | 0.6845 | 0.7800 | 1.3284 | 1.2616 | 1.3988 | 0.7915 | 0.7639 | 0.8200 |
| Least deprived (10) | 0.7634 | 0.7180 | 0.8118 | 1.3684 | 1.3029 | 1.4371 | 0.7344 | 0.7095 | 0.7603 |
| *Adjusted** | | | | | | | | | |
| Most deprived (1) | Reference | | | Reference | | | Reference | | |
| Decile 2 | 0.9609 | 0.9084 | 1.0163 | 1.1139 | 1.0572 | 1.1736 | 0.9891 | 0.9575 | 1.0217 |
| Decile 3 | 0.9250 | 0.8677 | 0.9860 | 1.1360 | 1.0737 | 1.2019 | 0.9765 | 0.9422 | 1.0121 |
| Decile 4 | 0.9849 | 0.9239 | 1.0500 | 1.1805 | 1.1144 | 1.2506 | 1.0121 | 0.9756 | 1.0500 |
| Decile 5 | 0.9364 | 0.8769 | 1.0000 | 1.1334 | 1.0698 | 1.2008 | 1.0009 | 0.9649 | 1.0383 |
| Decile 6 | 0.8918 | 0.8327 | 0.9551 | 1.1800 | 1.1153 | 1.2485 | 0.9321 | 0.8966 | 0.9689 |
| Decile 7 | 0.9398 | 0.8814 | 1.0021 | 1.1884 | 1.1258 | 1.2544 | 0.9129 | 0.8796 | 0.9474 |
| Decile 8 | 0.9329 | 0.8779 | 0.9914 | 1.1719 | 1.1130 | 1.2339 | 0.9416 | 0.9094 | 0.9750 |

| | | | | | | | | | |
|---|---|---|---|---|---|---|---|---|---|
| Decile 9 | 0.8518 | 0.7980 | 0.9091 | 1.2620 | 1.1991 | 1.3282 | 0.9241 | 0.8912 | 0.9582 |
| Least deprived (10) | 0.8925 | 0.8392 | 0.9493 | 1.2659 | 1.2056 | 1.3292 | 0.8804 | 0.8495 | 0.9123 |

Definitions: HR = Hazard Ratio, LCI = 95% Lower Confidence Interval, UCI = 95% Upper Confidence Interval

* Adjusted for age (10-year age bands), sex, ethnicity, long-term illness, number of tests in previous month, time varying vaccination status (with an interaction to time), previous infection status (and interaction to time), and 2019 Index of Multiple Deprivation score

**Table E: Results for a Cox regression predicting time to a registered positive test by deprivation decile for individuals who had received a flu vaccination within a year of the time period start date (as a proxy for healthcare engaged).**

| | **Delta (3rd June - 1st Sept 2021)** | | | **Delta (1st Sept - 27th Nov 2021)** | | | **Omicron (13th Dec - 2nd Feb2022)** | | |
|---|---|---|---|---|---|---|---|---|---|
| | **HR** | **LCI** | **UCI** | **HR** | **LCI** | **UCI** | **HR** | **LCI** | **UCI** |
| *Unadjusted* | | | | | | | | | |
| Most deprived (1) | Reference | | | Reference | | | Reference | | |
| Decile 2 | 0.9004 | 0.8570 | 0.9460 | 1.1614 | 1.1176 | 1.2070 | 1.0346 | 1.0020 | 1.0683 |
| Decile 3 | 0.8629 | 0.8168 | 0.9116 | 1.1747 | 1.1267 | 1.2246 | 1.0413 | 1.0056 | 1.0781 |
| Decile 4 | 0.9151 | 0.8651 | 0.9680 | 1.2129 | 1.1620 | 1.2661 | 1.0163 | 0.9796 | 1.0544 |
| Decile 5 | 0.8332 | 0.7875 | 0.8816 | 1.1427 | 1.0951 | 1.1925 | 1.0348 | 0.9988 | 1.0721 |
| Decile 6 | 0.7645 | 0.7217 | 0.8099 | 1.2144 | 1.1653 | 1.2655 | 0.9827 | 0.9482 | 1.0184 |
| Decile 7 | 0.7489 | 0.7092 | 0.7909 | 1.1482 | 1.1034 | 1.1948 | 0.9514 | 0.9195 | 0.9844 |
| Decile 8 | 0.7958 | 0.7564 | 0.8372 | 1.2331 | 1.1880 | 1.2799 | 0.9739 | 0.9430 | 1.0058 |
| Decile 9 | 0.6943 | 0.6576 | 0.7332 | 1.3341 | 1.2856 | 1.3844 | 0.9550 | 0.9239 | 0.9871 |
| Least deprived (10) | 0.6664 | 0.6322 | 0.7024 | 1.3712 | 1.3235 | 1.4206 | 0.9226 | 0.8935 | 0.9526 |
| *Adjusted** | | | | | | | | | |
| Most deprived (1) | Reference | | | Reference | | | Reference | | |
| Decile 2 | 0.9392 | 0.8937 | 0.9871 | 1.2106 | 1.1652 | 1.2577 | 1.0488 | 1.0154 | 1.0832 |
| Decile 3 | 0.9221 | 0.8725 | 0.9746 | 1.2485 | 1.1980 | 1.3011 | 1.0655 | 1.0286 | 1.1037 |
| Decile 4 | 1.0028 | 0.9476 | 1.0613 | 1.3338 | 1.2784 | 1.3918 | 1.0633 | 1.0244 | 1.1037 |
| Decile 5 | 0.9374 | 0.8855 | 0.9924 | 1.2878 | 1.2346 | 1.3434 | 1.0938 | 1.0551 | 1.1339 |
| Decile 6 | 0.8715 | 0.8222 | 0.9238 | 1.3767 | 1.3215 | 1.4341 | 1.0511 | 1.0135 | 1.0900 |

| | | | | | | | | | |
|---|---|---|---|---|---|---|---|---|---|
| Decile 7 | 0.8714 | 0.8245 | 0.9209 | 1.3257 | 1.2743 | 1.3791 | 1.0319 | 0.9965 | 1.0684 |
| Decile 8 | 0.9279 | 0.8814 | 0.9770 | 1.4323 | 1.3802 | 1.4864 | 1.0599 | 1.0255 | 1.0955 |
| Decile 9 | 0.8121 | 0.7685 | 0.8583 | 1.5363 | 1.4806 | 1.5941 | 1.0438 | 1.0089 | 1.0799 |
| Least deprived (10) | 0.7759 | 0.7353 | 0.8187 | 1.5405 | 1.4868 | 1.5962 | 1.0061 | 0.9734 | 1.0399 |

Definitions: HR = Hazard Ratio, LCI = 95% Lower Confidence Interval, UCI = 95% Upper Confidence Interval

* Adjusted for age (10-year age bands), sex, ethnicity, long-term illness, number of tests in previous month, time varying vaccination status (with an interaction to time), previous infection status (and interaction to time), and 2019 Index of Multiple Deprivation score

**Table F: Results for a Cox regression predicting time to a registered positive test for all residents in Cheshire and Merseyside.**

| | Delta (3rd June - 1st Sept 2021) | | | Delta (1st Sept - 27th Nov 2021) | | | Omicron (13th Dec – 28th Feb 2022) | | |
|---|---|---|---|---|---|---|---|---|---|
| | HR | LCI | UCI | HR | LCI | UCI | HR | LCI | UCI |
| *Unadjusted* | | | | | | | | | |
| Unvaccinated | Reference | | | Reference | | | Reference | | |
| 1 dose | 0.7395 | 0.7257 | 0.7536 | 0.5135 | 0.4992 | 0.5283 | 1.6952 | 1.6582 | 1.7330 |
| 2 doses | 0.4486 | 0.4411 | 0.4561 | 0.7058 | 0.6972 | 0.7146 | 2.4364 | 2.4064 | 2.4667 |
| 3 doses | 0.5793 | 0.4778 | 0.7023 | 0.5816 | 0.5162 | 0.6553 | 1.1328 | 1.1179 | 1.1478 |
| Previous infection | 0.2370 | 0.2236 | 0.2513 | 0.1493 | 0.1419 | 0.1572 | 0.8050 | 0.7928 | 0.8175 |
| Deprivation Score | 1.0031 | 1.0028 | 1.0034 | 0.9943 | 0.9940 | 0.9946 | 0.9994 | 0.9991 | 0.9996 |
| *Adjusted** | | | | | | | | | |
| Unvaccinated | Reference | | | Reference | | | Reference | | |
| 1 dose | 0.8250 | 0.8070 | 0.8434 | 0.8206 | 0.7957 | 0.8463 | 1.4238 | 1.3906 | 1.4579 |
| 2 doses | 0.7432 | 0.7275 | 0.7592 | 1.1416 | 1.1204 | 1.1632 | 2.1293 | 2.0981 | 2.1609 |
| 3 doses | 0.8262 | 0.6800 | 1.0037 | 0.9852 | 0.8773 | 1.1064 | 1.6127 | 1.5844 | 1.6415 |
| Previous infection | 0.2001 | 0.1886 | 0.2124 | 0.1300 | 0.1235 | 0.1369 | 0.6278 | 0.6181 | 0.6376 |
| Deprivation Score | 0.9998 | 0.9995 | 1.0001 | 0.9934 | 0.9932 | 0.9937 | 0.9985 | 0.9983 | 0.9987 |

Definitions: HR = Hazard Ratio, LCI = 95% Lower Confidence Interval, UCI = 95% Upper Confidence Interval

Note: Deprivation score is numerical, with increasing values representing higher levels of deprivation

* Adjusted for age (10-year age bands), sex, ethnicity, long-term illness, number of tests in previous month, time varying vaccination status (with an interaction to time), previous infection status (and interaction to time), and 2019 Index of Multiple Deprivation score

**Table G: Results for a Cox regression predicting time to a registered positive test for people aged 65 years and over who had received a flu vaccination within a year of the time period start date (as a proxy for healthcare engaged).**

| | Delta (3rd June - 1st Sept 2021) | | | Delta (1st Sept - 27th Nov 2021) | | | Omicron (13th Dec - 2nd Feb 2022) | | |
|---|---|---|---|---|---|---|---|---|---|
| | **HR** | **LCI** | **UCI** | **HR** | **LCI** | **UCI** | **HR** | **LCI** | **UCI** |
| *Unadjusted* | | | | | | | | | |
| Unvaccinated | Reference | | | Reference | | | Reference | | |
| 1 dose | 0.3643 | 0.2442 | 0.5435 | 1.0196 | 0.6243 | 1.6653 | 1.2246 | 1.1274 | 1.3301 |
| 2 doses | 0.4078 | 0.3160 | 0.5264 | 1.4593 | 1.0517 | 2.0248 | 1.4305 | 1.3583 | 1.5064 |
| 3 doses | 0.5008 | 0.2923 | 0.8580 | 0.3964 | 0.2582 | 0.6088 | 0.7568 | 0.7271 | 0.7878 |
| Previous infection | 0.2710 | 0.2035 | 0.3608 | 0.1484 | 0.1106 | 0.1991 | 0.6116 | 0.5749 | 0.6507 |
| Deprivation Score | 1.0097 | 1.0083 | 1.0112 | 1.0011 | 0.9999 | 1.0022 | 1.0011 | 1.0003 | 1.0018 |
| *Adjusted** | | | | | | | | | |
| Unvaccinated | Reference | | | Reference | | | Reference | | |
| 1 dose | 0.3465 | 0.2323 | 0.5170 | 1.0498 | 0.6433 | 1.7132 | 0.9742 | 0.8867 | 1.0703 |
| 2 doses | 0.4488 | 0.3473 | 0.5799 | 1.3987 | 1.0080 | 1.9410 | 1.1899 | 1.0768 | 1.3150 |
| 3 doses | 0.5665 | 0.3305 | 0.9711 | 0.4208 | 0.2748 | 0.6444 | 0.9631 | 0.8729 | 1.0628 |
| Previous infection | 0.2299 | 0.1725 | 0.3064 | 0.1207 | 0.0904 | 0.1611 | 0.4668 | 0.4385 | 0.4969 |
| Deprivation Score | 1.0087 | 1.0072 | 1.0103 | 1.0009 | 0.9997 | 1.0020 | 0.9992 | 0.9984 | 1.0000 |

Definitions: HR = Hazard Ratio, LCI = 95% Lower Confidence Interval, UCI = 95% Upper Confidence Interval

Note: Deprivation score is numerical, with increasing values representing higher levels of deprivation

* Adjusted for age (10-year age bands), sex, ethnicity, long-term illness, number of tests in previous month, time varying vaccination status (with an interaction to time), previous infection status (and interaction to time), and 2019 Index of Multiple Deprivation score
